# A Tear-Based Approach for Rapid Identification of Bacterial Pathogens in Corneal Ulcers Using Nanopore Sequencing

**DOI:** 10.1101/2024.09.26.24314375

**Authors:** Mark Dibbs, Mitchelle Matesva, Despoina Theotoka, Christina Jayaraj, Beruk Metiku, Patrick Demkowicz, Jacob S. Heng, Yvonne Wang, Christine Y. Bakhoum, Jessica Chow, Mathieu F. Bakhoum

**Author notes:** denotes equal contributions (Mark Dibbs, Mitchelle Matesva). Meeting Presentation: N/A. Address for Reprints: Mathieu F. Bakhoum, MD, PhD, 300 George St. #8107B, New Haven, CT 06511.

## Abstract

**Purpose:** Corneal ulcers pose a significant threat to vision, with the need for prompt and precise pathogen identification being critical to effective treatment. This study assesses the efficacy of using next-generation portable sequencing (Nanopore Technology) to detect and identify bacterial pathogens directly from tear samples, providing a non-invasive alternative to traditional corneal scraping and culture, which are limited by high false-negative rates.

**Design:** Prospective observational study.

**Participants:** Ten participants diagnosed with corneal ulcers.

**Methods:** Tear samples were collected from the ocular surface using Schirmer strips. Corneal scrapings and cultures were performed as medically indicated. The 16S rRNA gene was amplified directly from the tear samples using polymerase chain reaction (PCR), and Nanopore sequencing was used for bacterial species identification and taxonomic classification. Comparative analysis was conducted to evaluate the concordance between Nanopore sequencing results and traditional culture methods.

**Main Outcome Measures:** Comparison of bacterial species detected via Nanopore sequencing with those identified through traditional culture methods.

**Results:** Bacterial DNA was identified in 8 of the 10 samples analyzed using the tear-based sequencing method. Notably, Nanopore sequencing accurately identified the causative bacteria in all 4 samples that exhibited bacterial growth on culture. Additionally, it detected bacterial pathogens in 2 of the 4 ulcers that did not show bacterial growth on culture. In 2 cases where cultures could not be obtained due to the small size of the ulcer, tear sequencing successfully identified bacterial species, highlighting potentially overlooked pathogens in corneal ulcers.

**Conclusions:** PCR amplification of 16S RNA directly from tears followed by Nanopore sequencing is an effective, non-invasive method to identify bacterial pathogens in corneal ulcers, offering non-inferior results to traditional culture methods. This technique not only allows for the detection of traditionally hard-to-culture organisms, providing immediate diagnostic value to guide treatment, but also enhances our understanding of the microbiological landscape of corneal ulcers, thereby informing more effective treatment strategies.

## Introduction

Corneal ulcers represent a significant ophthalmological emergency, often leading to irreversible corneal damage and visual impairment. Globally, microbial keratitis, which is the primary cause of corneal ulcers, is estimated to contribute to approximately 1.5 to 2 million cases of unilateral blindness annually.^1^ In the United States, bacterial infections are most frequently responsible for these conditions.^2^ Traditionally, the diagnosis of corneal ulcers has relied on corneal scraping to collect samples for culturing and identifying the causative organism. This method, while considered the gold standard, is invasive, time-consuming, and exhibits variable sensitivity (38-66%),^3^ largely dependent on the size of the ulcer and the type of organism. Culture results, which can take several days to weeks, often delay the initiation of appropriate antimicrobial therapies, thereby extending patient discomfort and increasing the risk of complications and vision loss.

In contrast, next-generation sequencing technologies, such as Nanopore sequencing, offer a compelling alternative for microbial identification. This portable sequencing technology enables real-time basecalling, which significantly accelerates the pathogen identification process. Prior studies have validated the efficacy of Nanopore sequencing in various clinical scenarios, demonstrating its capability not only to identify microbial agents but also to predict antibiotic resistance.^4-8^ However, traditional approaches still require DNA extraction – and in the case of corneal ulcers invasive corneal scrapings – which necessitates specialized equipment and skills.

Tears possess a robust antimicrobial defense system which is crucial for protecting the ocular surface from infections. Factors such as Immunoglobulin A, which neutralizes and prevents pathogen adhesion; lysozyme, an enzyme that breaks down bacterial cell walls; lactoferrin, which possesses direct bactericidal effects; and enzymes like secretory Phospholipase A2 and Beta-lysin, which disrupt bacterial membranes, all contribute to this protective mechanism.^9,10^ These antimicrobial agents not only protect the eye but also facilitate the lysis of microbial cells, leading to the release of microbial DNA directly into the tears.

Under this premise, we hypothesized that it would be possible to detect microbial DNA directly from tear samples obtained from eyes with corneal ulcers, thus eliminating the need for corneal scrapings and DNA extraction. This would provide a non-invasive, efficient method for diagnosing ocular surface infections. Here, we conducted a prospective study on individuals presenting with bacterial corneal ulcers to compare the diagnostic sensitivity and specificity of the tear-based sequencing method to the traditional culture method.

## Methods

### Study Design and Subjects Enrollment

This is a prospective observational cohort study that was conducted in accordance with the principles of the Declaration of Helsinki and was approved by the Institutional Review Board of Yale University. Informed consent was obtained from all patients. Data on demographics, medical history, and medication use were collected from the patients’ medical records at the time of enrollment. Statistical analysis was performed using IBM SPSS Statistics software (version 29).

Participants included in the study were adults aged 18 years and older who had a clinical diagnosis of microbial keratitis confirmed by an ophthalmologist. Eligible individuals presented with symptoms consistent with corneal ulcers, such as eye redness, pain, and visual impairment, and were able to provide informed consent. Exclusion criteria included individuals with a history of corneal transplantation within the prior 6 months. We also excluded those diagnosed with viral or fungal keratitis based on preliminary clinical assessments, culture results, or prior medical records.

### Tear Collection

Tear samples were collected from the inferior fornix using a sterile Schirmer strip, avoiding contact with the eyelashes. The strip was subsequently placed in a microcentrifuge tube. Tears were collected following centrifugation for 2 minutes at 5000 RPM and stored at -80ºC.

### Corneal Scraping

Corneal scraping was performed as part of the standard of care for patients presenting with symptoms indicative of microbial keratitis, in accordance with established clinical guidelines. The decision to perform corneal scraping was made at the discretion of the treating physician based on the severity of the ulcer, size, location, and clinical presentation of the infection. The procedure was carried out using a slit lamp for magnification. The patient’s eye was anesthetized with a topical anesthetic to ensure comfort during the procedure. Using a sterile blade, a small sample of the epithelial tissue was carefully removed from the ulcer. The collected samples were immediately placed in appropriate culture media or transport media to preserve microbial viability and transported to the laboratory for prompt culturing and analysis.

### Nanopore 16S rRNA Amplification and Sequencing

Amplification and sequencing of the 16S rRNA gene, which is universal across bacterial species, were performed using the Oxford Nanopore Technology 16S Barcoding Kit 1-24 (SQK-16S024). The 16S rRNA gene was amplified using the LongAmp Hot Start Taq 2X Master Mix (New England Biolabs, Massachusetts, USA). One uL of the tear sample was added to the master mix and barcoded primers in a reaction mix volume of 50 uL. Polymerase chain reaction (PCR) was performed at the following settings: an initial denaturation step at 95ºC for 1 minute, followed by 50 cycles at 95ºC for 20 seconds, 55ºC for 30 seconds, and 65ºC for 2 minutes, and a final extension step at 65ºC for 5 minutes. The rest of the library preparation was performed using the recommended protocol for ONT kit SQK-16S024 for compatibility with the Flongle flow cell. PCR products were purified using AMPure XP Beads (Beckman Coulter, USA) following the Nanopore protocol. Sequencing runs were performed using the R9.4.1 Flongle flow cell (FLO-FLG001, Oxford Nanopore Technologies) on a MinION Mk1B Nanopore sequencer. Sequencing was allowed to proceed until a plateau of reads was achieved, which generally took between 4 and 12 hours.

### Analysis and Bacterial Identification

Basecalling was performed using the built-in Guppy basecaller on MinKnow, which translates the raw signal data from the Nanopore sequencer into nucleotide sequences. For species identification, the cloud-based EPI2ME FASTQ16S pipeline (v2023.04.21) provided by Oxford Nanopore Technology was utilized. The pipeline classifies the results from each sequencing run according to the NCBI 16S rRNA gene BLAST database. A minimum *qscore* of 7 was set, which indicates the quality threshold for basecalling accuracy; a higher *qscore* represents a higher confidence in the accuracy of the nucleotide base calls. In cases where multiple reads were identified, the determination of the causative organism was based on the highest read count.

## Results

### Demographics of study participants and clinical presentation

There were 10 subjects who were included after meeting the study criteria. The mean age was 50.2 years. There were 6 males and 4 females. The ulcer was present in the left eye of 5 subjects and right eye of 5 subjects. Patient demographics are summarized in **Table 1**.

**Table 1.**
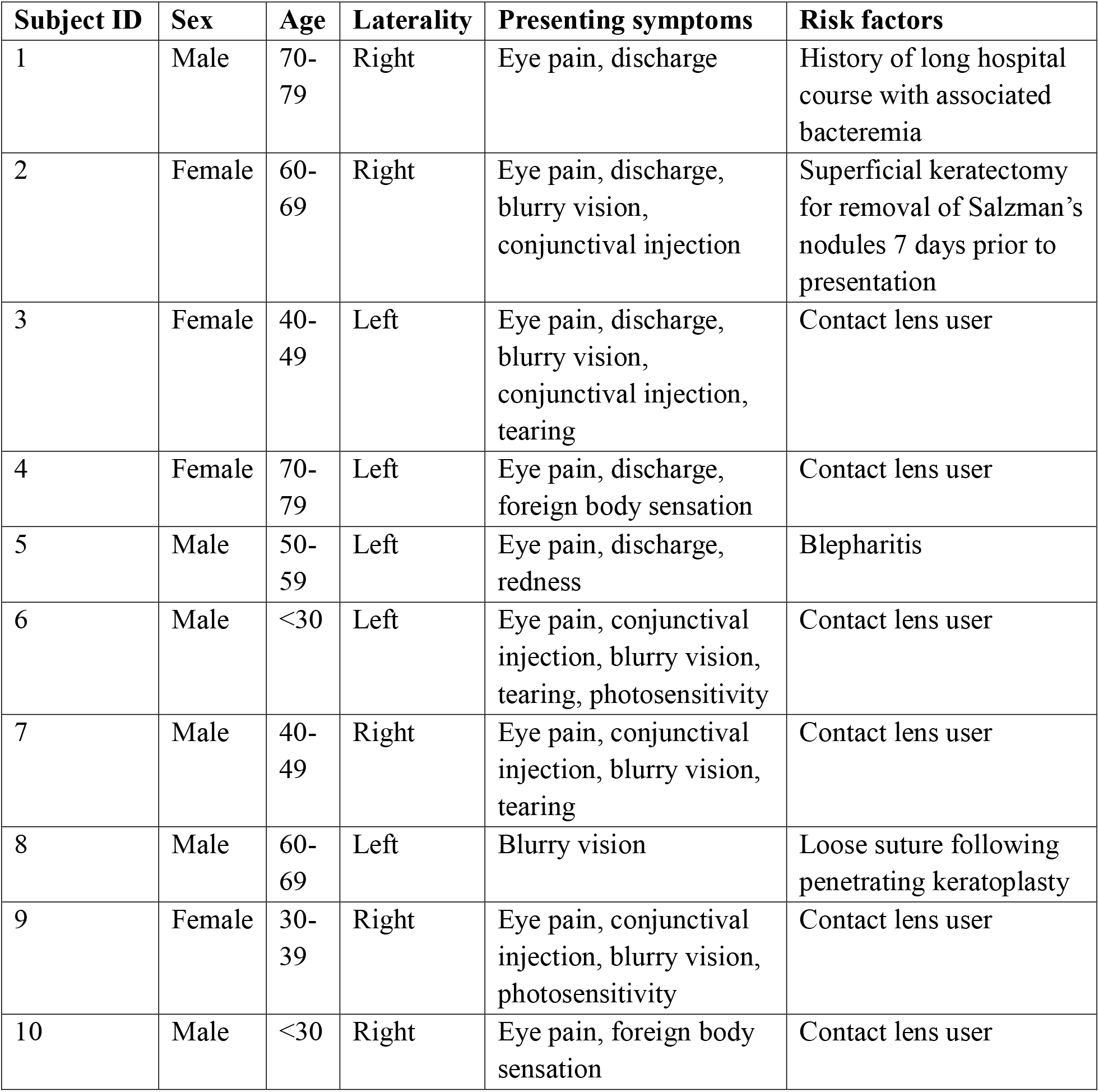
Demographics and Clinical Characteristics of the Study Subjects.

| **Subject ID** | **Sex** | **Age** | **Laterality** | **Presenting symptoms** | **Risk factors** |
| --- | --- | --- | --- | --- | --- |
| 1 | Male | 70-79 | Right | Eye pain, discharge | History of long hospital course with associated bacteremia |
| 2 | Female | 60-69 | Right | Eye pain, discharge, blurry vision, conjunctival injection | Superficial keratectomy for removal of Salzmann's nodules 7 days prior to presentation |
| 3 | Female | 40-49 | Left | Eye pain, discharge, blurry vision, conjunctival injection, tearing | Contact lens user |
| 4 | Female | 70-79 | Left | Eye pain, discharge, foreign body sensation | Contact lens user |
| 5 | Male | 50-59 | Left | Eye pain, discharge, redness | Blepharitis |
| 6 | Male | <30 | Left | Eye pain, conjunctival injection, blurry vision, tearing, photosensitivity | Contact lens user |
| 7 | Male | 40-49 | Right | Eye pain, conjunctival injection, blurry vision, tearing | Contact lens user |
| 8 | Male | 60-69 | Left | Blurry vision | Loose suture following penetrating keratoplasty |
| 9 | Female | 30-39 | Right | Eye pain, conjunctival injection, blurry vision, photosensitivity | Contact lens user |
| 10 | Male | <30 | Right | Eye pain, foreign body sensation | Contact lens user |

Symptoms at presentation included blurred vision, pain, pink eye, foreign body sensation, discharge, tearing, and sensitivity to light. Time of symptom onset prior to presentation ranged from 1 to 14 days with a mean of 4.7 days. Risk factors for the development of corneal ulcer included use of soft contact lenses (n = 6), history of long hospitalization with associated bacteremia (n = 1), blepharitis (n = 1), status post superficial keratectomy 7 days prior to diagnosis (n = 1), and loose suture in a patient with a penetrating keratoplasty (n = 1). Best corrected or pinhole visual acuity at presentation ranged from 0 to 2.40 LogMAR with a mean of 1.07 LogMAR, and a standard deviation of 0.88 LogMAR.

On examination, 9 patients presented with a single corneal infiltrate while one patient had 2 infiltrates of comparable size. Corneal ulcers were centrally located in 50% of the cases, with the remainder being peripheral. The average size of infiltrates measured 2.09 mm vertically (range, 1 mm to 5 mm) and 2.20 mm horizontally (range, 1 mm to 5 mm). No ulcers extended to the sclera or limbus. Two patients developed hypopyon.

Eight subjects underwent corneal scraping for culture, while 2 had ulcers considered too small for scraping. A total of 10 tear samples were collected.

### Summary of culture results

Of the 8 subjects who underwent corneal scraping and culture, 4 exhibited bacterial growth on culture. The bacteria identified included coagulase negative *Staphylococcus* in broth (Subject 1), *Staphylococcus aureus* (Subject 2), and *Pseudomonas aeruginosa* (Subjects 3 and 4), all identified one day post-diagnosis. The remaining 4 subjects showed no bacterial growth on culture media including blood agar, chocolate agar, and thioglycolate broth, which were monitored for growth for 5 days. Results from culture are listed in **Table 2**.

**Table 2.** Summary of Culture Results and Nanopore Sequencing Results.

| **Subject ID** | **Culture results** | **Nanopore sequencing results** | **Concordance** |
| --- | --- | --- | --- |
| 1 | Coagulase-negative<br>*Staphylococcus* | *Staphylococcus*<br>*saccharolyticus* | Yes |
| 2 | *Staphylococcus*<br>*aureus* | *Staphylococcus*<br>*roterodami* | Yes |
| 3 | *Pseudomonas*<br>*aeruginosa* | *Pseudomonas*<br>*aeruginosa* | Yes |
| 4 | *Pseudomonas*<br>*aeruginosa* | *Pseudomonas*<br>*aeruginosa* | Yes |
| 5 | No growth | *Staphylococcus*<br>*saccharolyticus* | No |
| 6 | No growth | *Cutibacterium acnes* | No |
| 7 | No growth | no bacteria detected | Yes |
| 8 | No growth | no bacteria detected | Yes |
| 9 | Too small to culture | *Staphylococcus*<br>*saccharolyticus* | Not applicable |
| 10 | Too small to culture | *Kocuria rhizophila* | Not applicable |

### Nanopore sequencing results

For the 4 subjects whose ulcer scrapings exhibited growth on culture, Nanopore sequencing identified the predominant bacterial pathogens in each tear sample as follows: *Staphylococcus saccharolyticus, Staphylococcus roterodami, Pseudomonas aeruginosa*, and again *Pseudomonas aeruginosa*, respectively. *Staphylococcus saccharolyticus* is coagulase negative,^11^ and *Staphylococcus roterodami* is within the *Staphylococcus aureus* complex.^12^

Among the 4 subjects whose ulcers did not exhibit growth on culture, Nanopore sequencing detected no bacteria in 2 cases, while identifying *Staphylococcus saccharolyticus* and *Cutibacterium acnes* in the remaining 2.

For the 2 subjects with ulcers too small for culture, Nanopore sequencing identified *Staphyloccous saccharolyticus* in one and *Kocuria rhizophila* in the other. Results from Nanopore sequencing compared to results from culture are listed in **Table 2**. Detailed results featuring taxonomic classification for each case along with a synopsis of the clinical presentation are provided in **Figures S1-S8**.

## Discussion

Our findings demonstrate the efficacy of PCR amplification of the 16S gene directly from tears, followed by Nanopore sequencing, in identifying the causative bacterial agents of corneal ulcers. This method demonstrated high sensitivity, successfully detecting bacterial pathogens in all samples that were positive by traditional culture methods. Notably, it also identified bacterial DNA in cases where traditional cultures failed, suggesting that tear-based Nanopore sequencing may offer greater sensitivity compared to standard culture-based methods. This non-invasive approach eliminates the need for DNA extraction and delivers results within hours, which is crucial for the timely management of corneal ulcers.

Traditional culture-based diagnostics are often limited by high false-negative rates, which range between 32.6-79.4%.^13,14^ This limitation stems from several factors, including variability in sample collection techniques and the fastidious nature of certain pathogens that do not grow under standard culture conditions. In contrast, our tear-based sequencing approach, which relies on detecting bacterial DNA, should in theory circumvent these hurdles. Expectedly, in our study, this method identified bacterial reads in 2 of the 4 ulcers that did not exhibit growth on culture, indicating higher sensitivity than traditional methods. Specifically, Nanopore sequencing identified *Staphylococcus saccharolyticus* and *Cutibacterium acnes* as the predominant bacterial species in culture-negative ulcers, both of which are known to be associated with corneal ulcers or eye infections.^15,16^ Notably, *Cutibacterium acnes*, formerly known as *Propionibacterium acnes*, is considered a fastidious organism due to its stringent growth requirements.^17,18^

Moreover, the tear-based approach could be particularly useful in clinical settings where access to an ophthalmologist is not feasible or laboratory resources are limited, as it requires no specialized equipment or training for sample collection, compared with the traditional culture techniques which require corneal scrapings by an ophthalmologist using a slit-lamp. The sequencing method offers significant benefits because it is non-invasive, eliminating the discomfort and potential complications associated with corneal scrapings. Additionally, its ability to bypass DNA extraction helps deliver rapid results, meeting a critical clinical need for quick and accurate diagnostics to guide treatment.

Another advantage of this sequencing-based method is its ability to detect low-abundance pathogens and provide insights into the polymicrobial nature of corneal ulcers. In our study, causative organisms identified by Nanopore included pathogens commonly associated with corneal ulcers such as *Staphylococcus, Pseudomonas*, and *Cutibacterium acnes*. Notably, *Kocuria rhizophila* has not been previously reported as a cause of corneal ulcers. It belongs to the *Kocuria* genus, a group of gram-positive bacteria that, in rare cases, can cause infectious keratitis in immunocompromised patients.^19-21^ Nanopore sequencing was particularly effective at identifying causative bacterial agents in corneal ulcers that were too small for conventional scraping and culture, highlighting its utility in detecting the polymicrobial etiology of these infections. Of the 8 eyes that yielded positive results on Nanopore, 7 showed reads from more than one bacterial genus. This finding aligns with previous reports on the polymicrobial nature of infection-induced corneal ulcers. For instance, in one study, among 81 corneal ulcers analyzed via culture, 43% yielded more than one bacterial organism.^22^ Other studies have estimated that between 1.9% and 25% of corneal ulcers are polymicrobial in nature.^23-25^ Our findings suggest that polymicrobial infections may be more common than previously recognized, possibly due to the enhanced detection capabilities of this new method. These findings also indicate that in corneal ulcers, a favorable environment may be conducive to the growth of multiple bacterial species.

This study is limited by its small sample size and the fact that it was conducted at a single center, which may restrict the generalizability of the findings. Additionally, the risk of contamination from eyelid skin or during the tear collection and sequencing processes cannot be completely ruled out. Some detected bacterial species might represent contaminants from the normal ocular flora.^26-29^ Furthermore, while Nanopore sequencing demonstrated high sensitivity and specificity, its accuracy needs to be validated across a broader spectrum of bacterial, fungal, and viral pathogens.^30^ Future research should also explore integrating this technology with antibiotic susceptibility testing to enhance its clinical utility and inform more targeted treatment strategies. Expanding the study to include more participants and multiple sites, along with testing for fungal and viral pathogens and conducting real-time antimicrobial resistance profiling, are next steps to build on these novel findings.

## Data Availability

All data produced in the present study are available upon reasonable request to the authors.

## Abbreviations and Acronyms

PCR: polymerase chain reaction

