## Supplementary Figure S1 for "A Tear-Based Approach for Rapid Identification of Bacterial Pathogens in Corneal Ulcers Using Nanopore Sequencing"

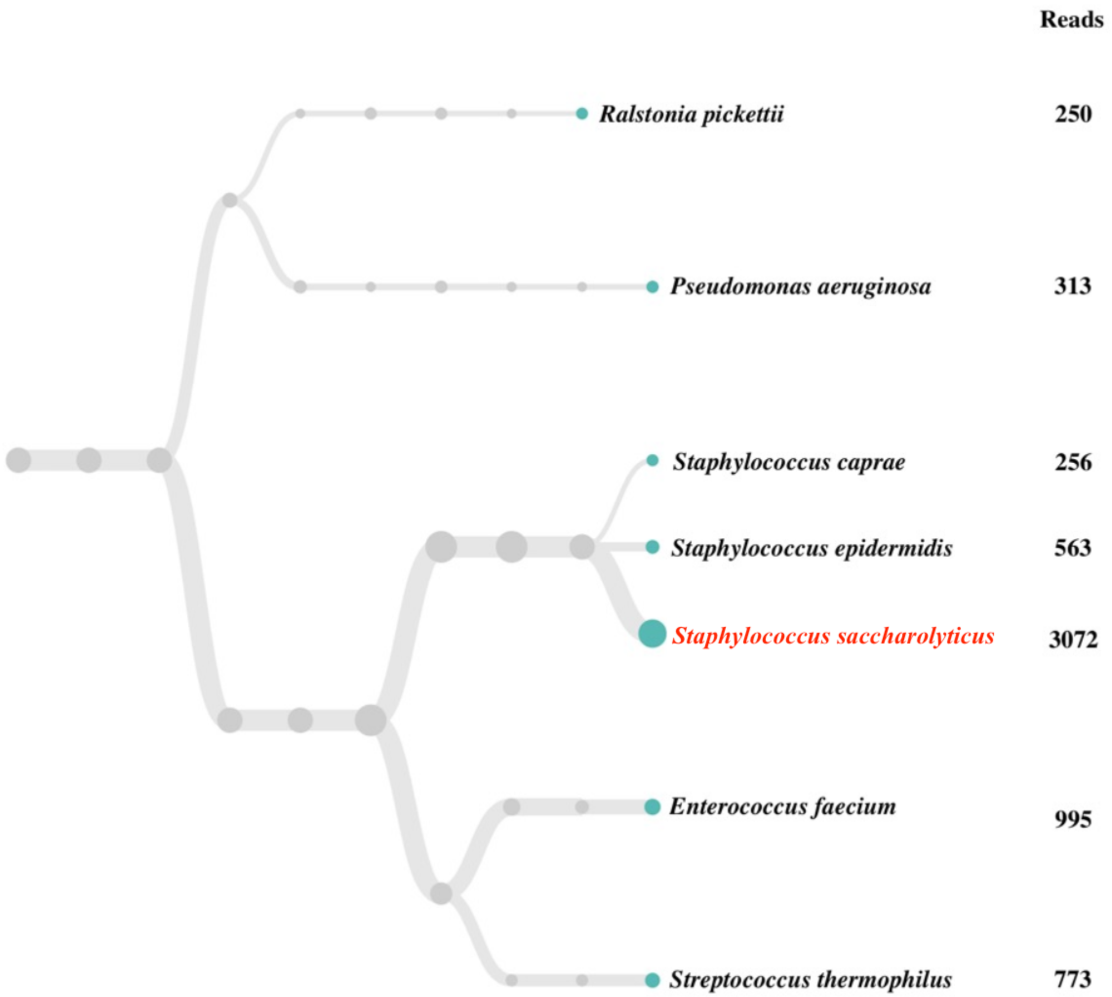


**Figure S1.** Taxonomic tree of bacterial species identified from the right eye of Subject 1. He presented with eye pain and discharge after experiencing a prolonged hospitalization complicated by bacteremia. The bacterial species with the greatest number of reads was *Staphylococcus saccharolyticus*, accounting for 42.1% of all classified reads.
