## Supplementary Figure S2 for "A Tear-Based Approach for Rapid Identification of Bacterial Pathogens in Corneal Ulcers Using Nanopore Sequencing"

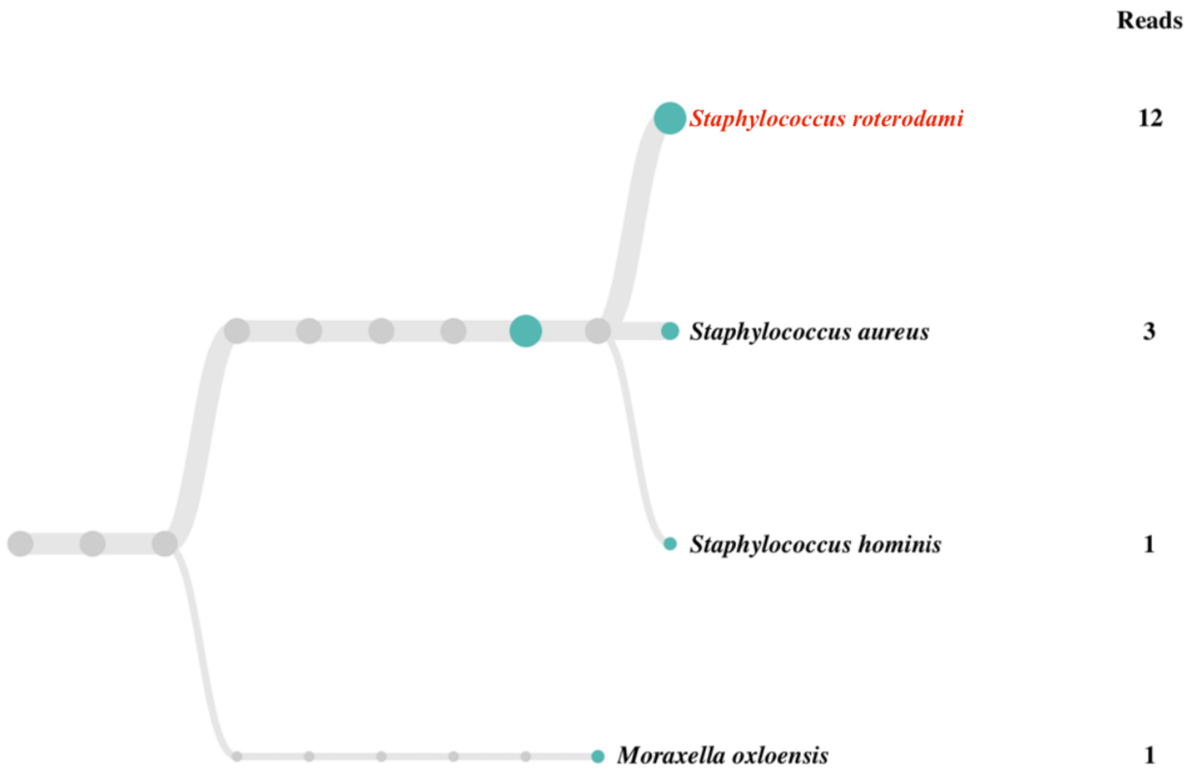


**Figure S2.** Taxonomic tree of bacterial species identified from the right eye of Subject 2. She presented with eye pain, discharge, blurry vision, and pink eye, and had a superficial keratectomy for removal of Salzmann's nodules 7 days prior to presentation. The bacterial species with the greatest number of reads was *Staphylococcus roterodami*, accounting for 66.7% of all classified reads.
