## Supplementary Figure S3 for "A Tear-Based Approach for Rapid Identification of Bacterial Pathogens in Corneal Ulcers Using Nanopore Sequencing"

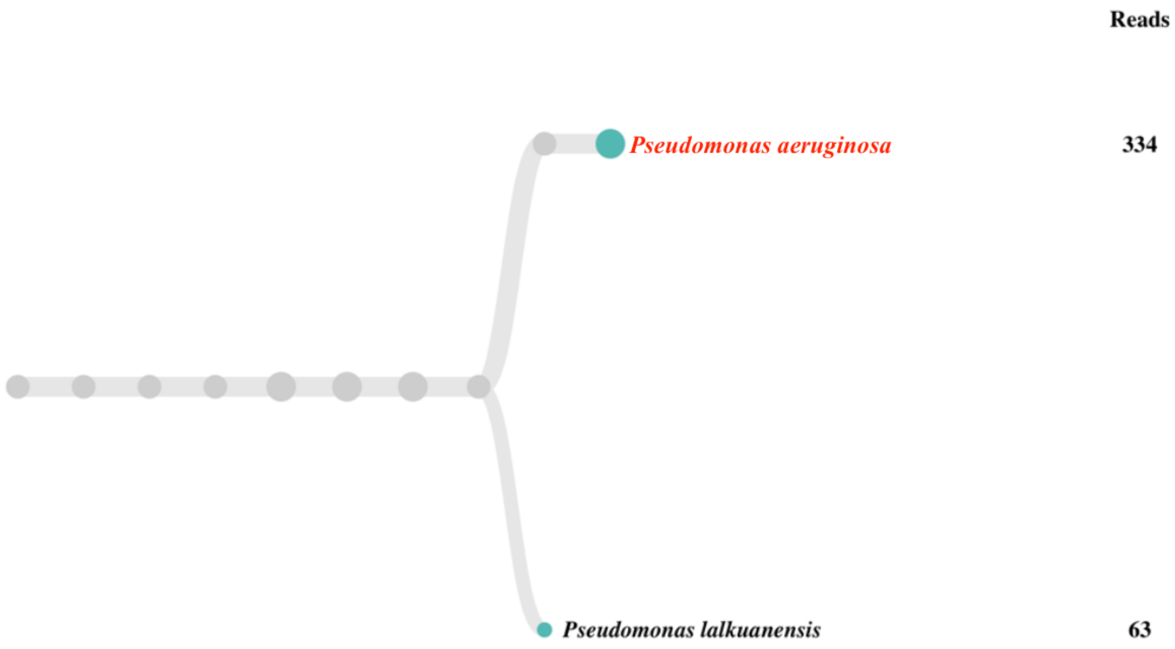


**Figure S3.** Taxonomic tree of bacterial species identified from the left eye of Subject 3. She was a contact lens user who presented with eye pain, discharge, blurry vision, pink eye, and tearing. The bacterial species with the greatest number of reads was *Pseudomonas aeruginosa*, accounting for 72.0% of all classified reads.
