## Supplementary Figure S4 for "A Tear-Based Approach for Rapid Identification of Bacterial Pathogens in Corneal Ulcers Using Nanopore Sequencing"

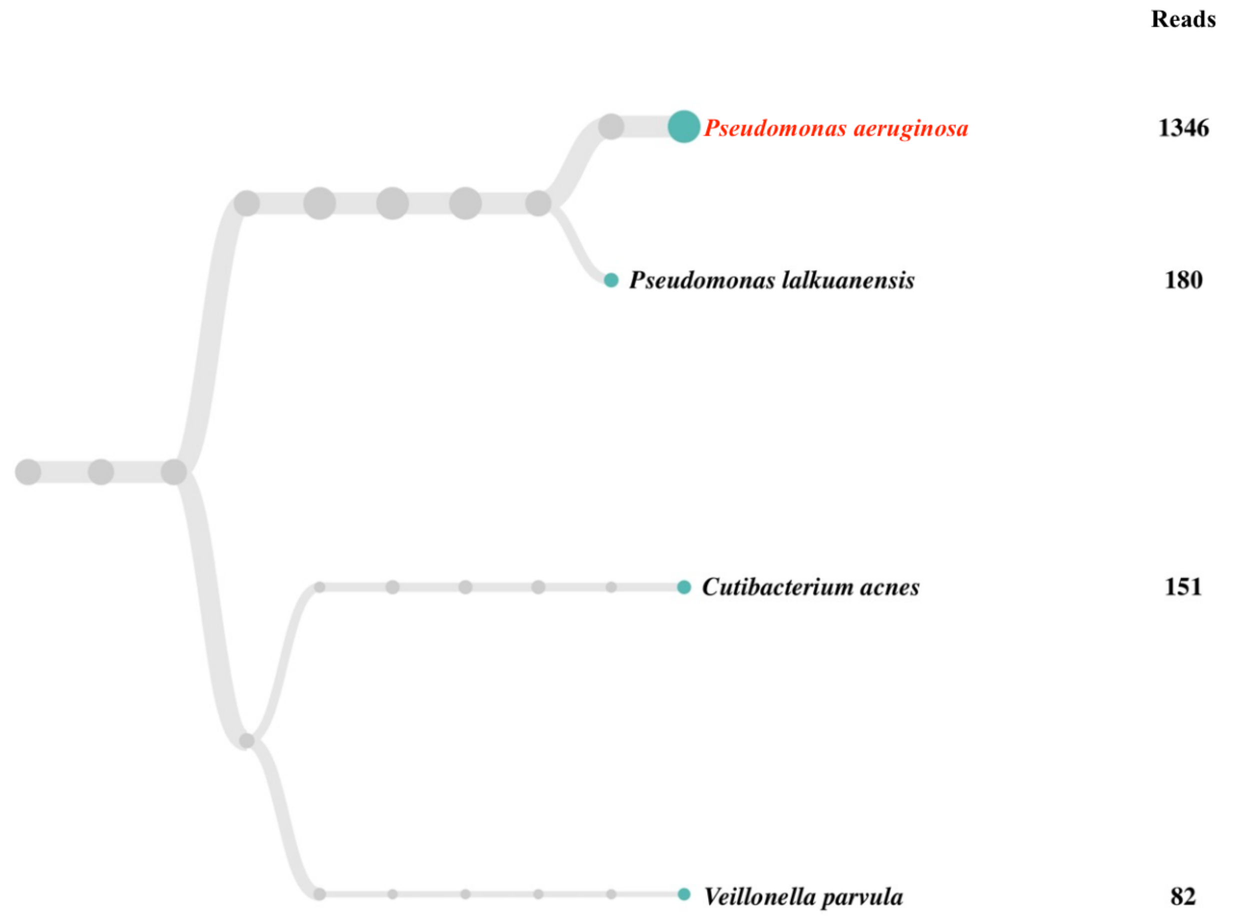


**Figure S4.** Taxonomic tree of bacterial species from the left eye of Subject 4. She was a contact lens user who presented with eye pain, discharge, and foreign body sensation. The bacterial species with the greatest number of reads was *Pseudomonas aeruginosa*, accounting for 62.1% of all classified reads.
