## Supplementary Figure S5 for "A Tear-Based Approach for Rapid Identification of Bacterial Pathogens in Corneal Ulcers Using Nanopore Sequencing"

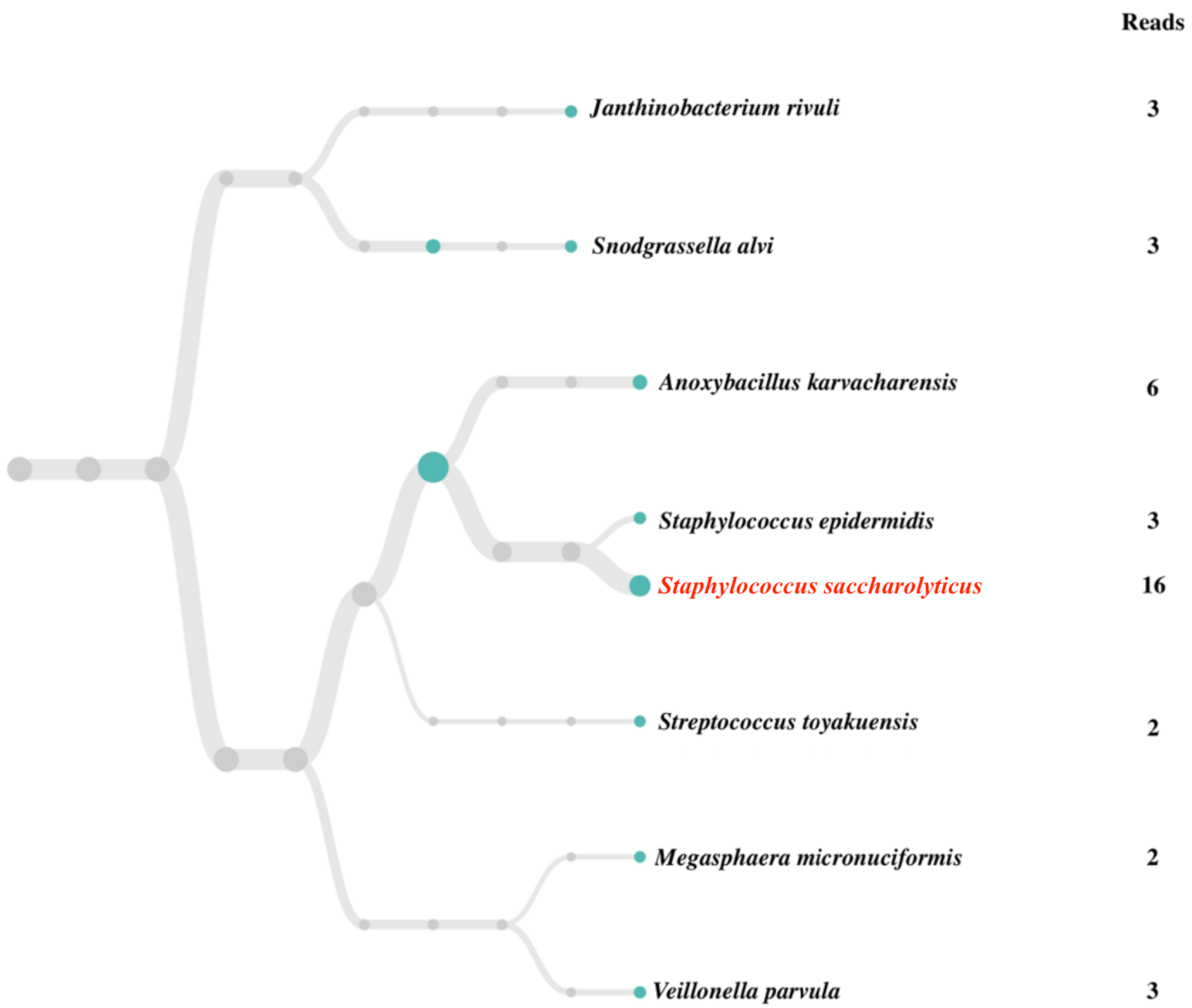


**Figure S5.** Taxonomic tree of bacterial species identified from the left eye of Subject 5. He had blepharitis and presented with eye pain, discharge, and redness. The bacterial species with the greatest number of reads was *Staphylococcus saccharolyticus*, accounting for 26.7% of all classified reads.
