## Supplementary Figure S6 for "A Tear-Based Approach for Rapid Identification of Bacterial Pathogens in Corneal Ulcers Using Nanopore Sequencing"

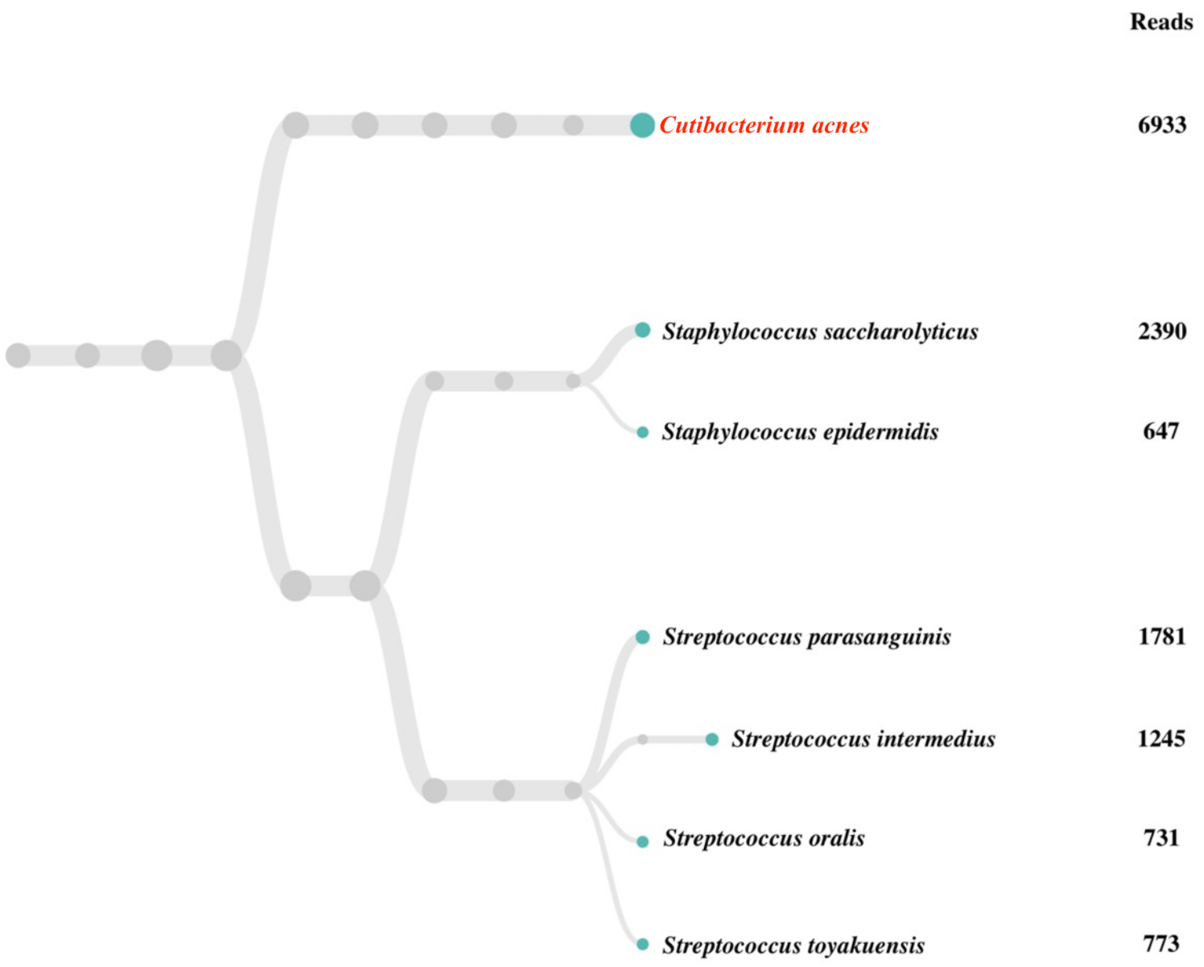


**Figure S6.** Taxonomic tree of bacterial species identified from the left eye of Subject 6. He was a contact lens user who presented with eye pain, pink eye, blurry vision, tearing, and photosensitivity. The bacterial species with the greatest number of reads was *Cutibacterium acnes*, accounting for 35.4% of all classified reads.
