## Supplementary Figure S7 for "A Tear-Based Approach for Rapid Identification of Bacterial Pathogens in Corneal Ulcers Using Nanopore Sequencing"

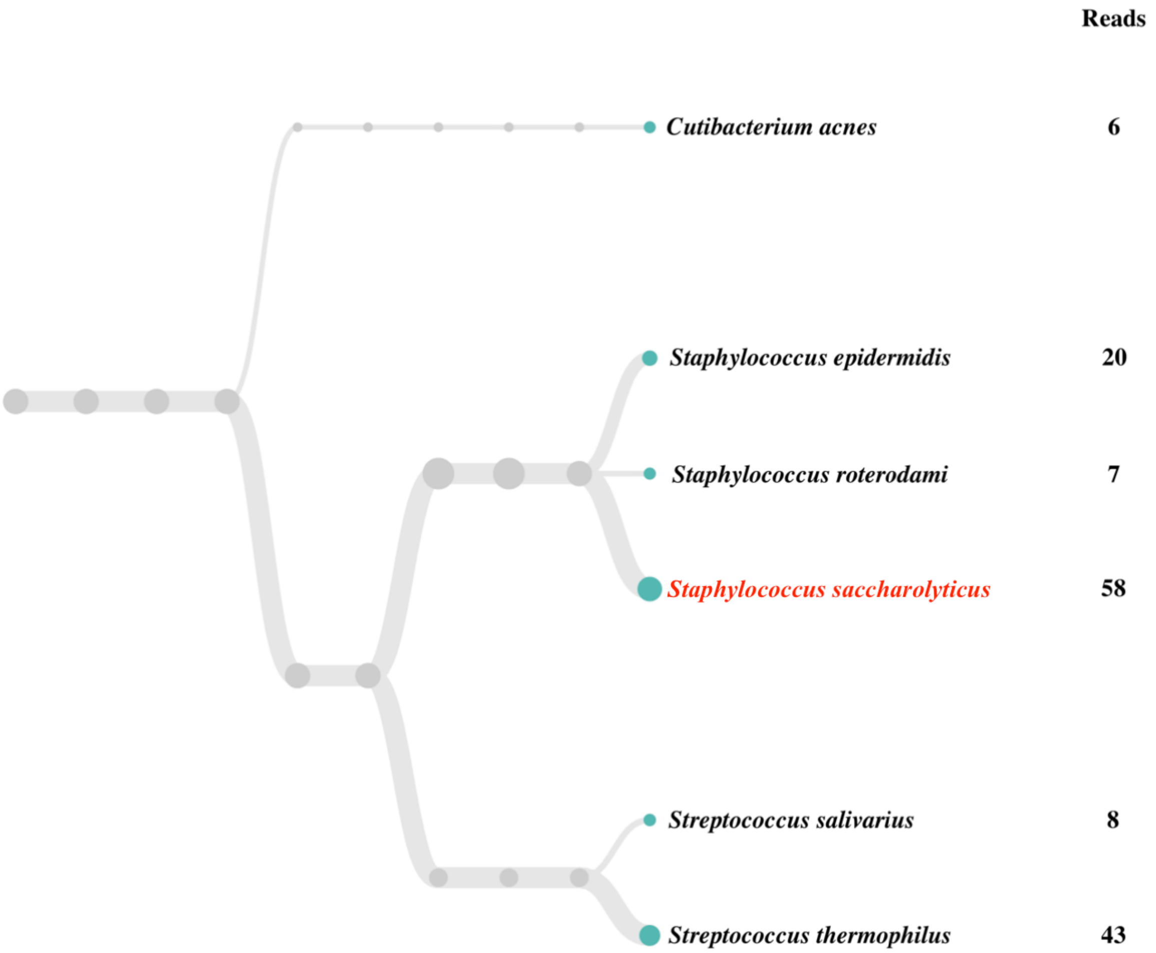


**Figure S7.** Taxonomic tree of bacterial species identified from the right eye of Subject 9. She was a contact lens user who presented with eye pain, pink eye, blurry vision, and photosensitivity. The bacterial species with the greatest number of reads was *Staphylococcus saccharolyticus*, accounting for 33.3% of all classified reads.
