## Supplementary Figure S8 for "A Tear-Based Approach for Rapid Identification of Bacterial Pathogens in Corneal Ulcers Using Nanopore Sequencing"

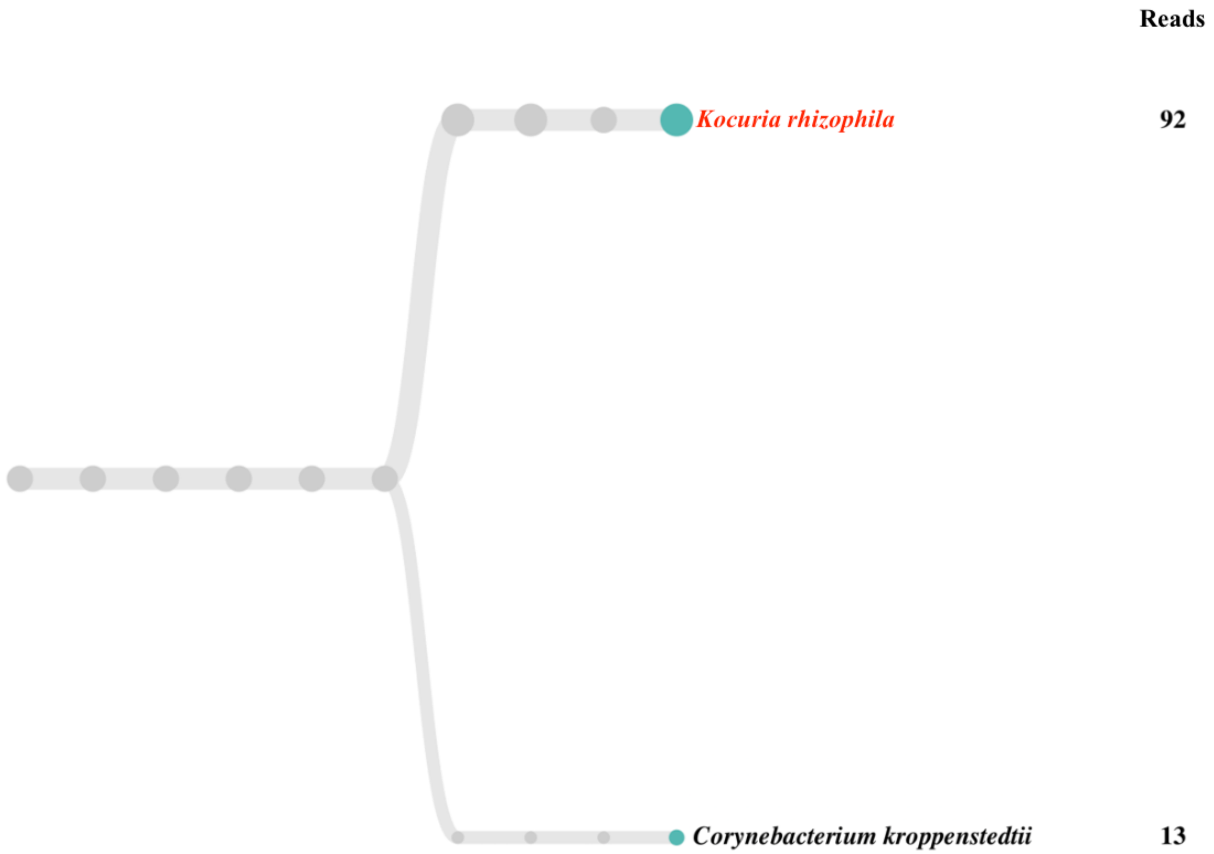


**Figure S8.** Taxonomic tree of bacterial species identified from the right eye of Subject 10. He was a contact lens user who presented with eye pain and foreign body sensation. The bacterial species with the greatest number of reads was *Kocuria rhizophila*, accounting for 78.0% of all classified reads.
